# Comprehensive assessment of humoral response after Pfizer BNT162b2 mRNA Covid-19 vaccination: a three-case series

**DOI:** 10.1101/2021.03.19.21253989

**Authors:** Elisa Danese, Martina Montagnana, Gian Luca Salvagno, Matteo Gelati, Denise Peserico, Laura Pighi, Simone De Nitto, Brandon M. Henry, Stefano Porru, Giuseppe Lippi

## Abstract

**Background:** Since universal vaccination is a pillar against coronavirus disease 2019 (COVID-19), monitoring anti-SARS-CoV-2 neutralizing antibodies is essential for deciphering post-vaccination immune response.

**Methods:** Three healthcare workers received 30 μg BNT162b2 mRNA Covid-19 Vaccine, followed by a second identical dose, 21 days afterwards. Venous blood was drawn at baseline and at serial intervals, up to 63 days afterwards, for assessing total immunoglobulins (Ig) anti-RBD (receptor binding domain), IgG anti-S1/S2, IgG anti-RBD, IgM anti-RBD, IgM anti-N/S1 and IgA anti-S1.

**Results:** All subjects were SARS-CoV-2 seronegative at baseline. Total Ig anti-RBD, IgG anti-S1/S2 and IgG anti-RBD levels increased between 91-368 folds until 21 days after the first vaccine dose, then reached a plateau. The levels raised further after the second dose (by ∼30-, ∼8- and ∼8-fold, respectively), peaking at day 35, but then slightly declining and stabilizing ∼50 days after the first dose. IgA anti-S1 levels increased between 7-11 days after the first dose, slightly declined before the second dose, after which levels augmented by ∼24-fold from baseline. The anti-RBD and anti-N/S1 IgM kinetics were similar to that of anti-S1 IgA, though displaying substantially weaker increases and modest peaks, only 4 to 7-fold higher than baseline. Highly significant inter-correlation was noted between total Ig anti-RBD, anti-S1/S2 and anti-RBD IgG (all r=0.99), whilst other anti-SARS-CoV-2 antibodies displayed lower, though still significant, correlations. Serum spike protein concentration was undetectable at all time points.

**Conclusions:** BNT162b2 mRNA vaccination generates a robust humoral immune response, especially involving IgG and IgA, magnified by the second vaccine dose.

## Introduction

Although social distancing, hand hygiene, widespread use of face masks, and contact tracing are essential measures for limiting the burden of severe acute respiratory syndrome coronavirus 2 (SARS-CoV-2) infections [1], the worldwide spread of coronavirus disease 2019 (COVID-19) appears nearly unstoppable, especially after the emergence of new and potentially more aggressive variants [2]. Several advancements made in the therapeutic management of patients with COVID-19 have considerably contributed to improve clinical outcomes, thus lowering SARS-CoV-2-related morbidity, disability and death [3]. Nonetheless, universal vaccination, paving the way to the generation of efficient and durable personal and herd immunity, is now regarded as the key pillar for mitigating the dramatic impact of SARS-CoV-2 on healthcare, society and economy [4]. Among the various types of COVID-19 vaccines in the advanced stage of development, or already approved for clinical use, those based on mRNA technology were the first approved and administered in many western countries due to some technical advantages, comprehensively summarized elsewhere [5]. Earlier phase III trials and real-world data demonstrate that these vaccines are effective to mitigate COVID-19 severity, and this is implicitly attributable to the generation of an efficient humoral and cellular immunity, in particular, the development of a robust anti-SARS-CoV-2 neutralizing antibody response [6-7]. Nonetheless, published data describing the early and comprehensive humoral immune response after mRNA COVID-19 vaccination in subjects not included in clinical trials is still lacking to the best of our current knowledge.

## Materials and Methods

This 3-case series was based on two female (44 and 39 years old) and one male (53 years old) healthcare workers, who received the first 30 μg BNT162b2 mRNA Covid-19 Vaccine dose (Comirnaty, Pfizer Inc, NY, USA) at the University Hospital of Verona (Italy) on January 7, 2021, followed by a second identical dose 21 days afterwards (January 28, 2021). Venous blood was collected in the morning by straight needle venipuncture into 5 mL evacuated blood tubes containing gel and clot activator (Vacutest, Kima, Padova, Italy) on theday before first dose administration, and then on 1, 4, 7, 11, 14, 21 (2 hours before second vaccination), 22, 25, 28, 35, 42, 49, 56 and 63 days afterwards. The study volunteers were not taking immunomodulatory drugs, whilst lifestyle, diet, level of physical activity remained unchanged throughout the study period. Blood samples were centrifuged at 1500xg for 15 min at room temperature; serum was separated and stored in two aliquots of ∼1 mL at −70°C. All serum aliquots were concomitantly thawed at the end of the study period, centrifuged and tested with six anti-SARS-CoV-2 immunoassays (Table 1), as well as with a manual enzyme linked immunoassay (ELISA) for quantification of serum SARS-CoV-2 spike concentration (COVID-19 Spike Protein ELISA Kit, Abcam, Cambridge, UK; measuring range: 2.7-2000 ng/mL, intra-assay imprecision: <10%). Previous evidence has been published that four of these six anti-SARS-CoV-2 immunoassays display good concordance with virus neutralization tests (Table 1) [8,9]. Molecular testing for detecting SARS-CoV-2 RNA (Seegene AllplexTM2019-nCoV Assay, Seegene, Seoul, South Korea) was also carried out on nasopharyngeal swabs 3 days before receiving the first vaccine dose, and then every ∼2 weeks afterwards, up till the end of the study. Cumulative results of antibodies testing were presented as mean ± standard deviation (SD), as ratio with baseline antibodies level (i.e., [time point value]/[baseline value and/or limit of detection]). Pearson’s test was used to test the correlation of levels of different antibody classes over time. Statistical analysis was conducted with Analyse-it (Analyse-it Software Ltd, Leeds, UK). The three subjects involved in this case series are three authors of the article (E.D., M.M. and G.L.), who underwent routine institutional laboratory monitoring by means of nasopharyngeal swabs and venous blood sampling throughout the study period, according to a protocol approved by the local Ethics Committee of Verona and Rovigo (2683CESC; February 16, 2021).

**Table 1.** Principal technical and analytical features of anti-SARS-CoV-2 (severe acute respiratory syndrome coronavirus 2) antibodies immunoassays used in this study.

| Test | Company | Analyzer | Principle | Ig class | Target | Cut-off | Agreement with VNT |
| --- | --- | --- | --- | --- | --- | --- | --- |
| Elecsys Anti-SARS-CoV-2 S | Roche | Cobas 8000 | CLIA | Total Ig | RBD | $\geq 0.80$ U/mL | N/A |
| VIDAS SARS-COV-2 IgM | BioMérieux | VIDAS | ELFA | IgM | RBD | $> 0.960$ OD | AUC vs. PRNT<sub>50</sub> $\geq 20$: 0.80 (95% CI, 0.72-0.88) [8] |
| TGS COVID-19 IgM | Technogenetics | IDS-iSYS | CLIA | IgM | Nucleocapsid/Spike (S1) | $\geq 1.0$ (index) | N/A |
| Anti-SARS-CoV-2 ELISA IgA | Euroimmun | Manual | ELISA | IgA | Spike (S1) | $\geq 1.1$ (ratio) | AUC vs. PRNT<sub>50</sub> $\geq 20$: 0.91 (95% CI, 0.85-0.97) [8] |
| ACCESS SARS-CoV-2 IgG II | Beckman Coulter | Access 2 | CLIA | IgG | RBD | $\geq 10$ U/mL | Correlation with NT<sub>50</sub>: 0.73-0.77 [9] |
| LIAISON SARS-CoV-2 TrimericS IgG | DiaSorin | LIAISON XL | CLIA | IgG | Spike (S1/S2) | $\geq 13$ U/mL | AUC vs. PRNT<sub>50</sub> $\geq 20$: 0.96 (95% CI, 0.94-0.99) [8] |

*AUC, area under the curve; CLIA, ChemiLuminescent ImmunoAssay; ELFA, Enzyme Linked Fluorescent Assay; ELISA, Enzyme Linked ImmunoSorbent Assay; Ig, Immunoglobulin; N, nucleocapsid; N/A, Not Available; OD, Optical Density, PRNT<sub>50</sub>, plaque reduction neutralization test by 50%; RBD, Receptor Binding Domain; VNT, Virus Neutralization Test.*

## Results

Molecular testing for detecting SARS-CoV-2 RNA repeatedly yielded negative results in all three study subjects. Moreover, no clinical sign or symptoms of COVID-19 occurred in any participant, such that ongoing (even asymptomatic) SARS-CoV-2 infection could be ruled out throughout the study period. No major side effects were recorded after administration of either vaccine dose. Mild pain at the injection site, lasting between 8-36 hours, was observed after both vaccine doses.

The cumulative results of this study are shown in Figure 1. In all the three subjects, who were SARS-CoV-2 seronegative at baseline, serum levels of all anti-SARS-CoV-2 antibodies started to increase between 7-11 days after the first vaccine dose, though displaying some different kinetics. Total Ig anti-RBD, IgG anti-S1/S2 and IgG anti-RBD increased gradually between 91-368 folds from day 11 after the first dose up to 21 days afterwards, when a plateau was reached. A further increase (of ∼30-fold for total Ig anti-RBD and ∼8-fold for both IgG anti-S1/S2 and IgG anti-RBD; Fig. 2) could be noted after the second vaccine dose, peaking 35 days after the first vaccine dose (Fig. 1a). After this time point, the levels of all these antibodies classes slightly declined, reaching a second plateau 50 days after the first vaccine dose, with values still higher than the first peak observed after the first vaccine dose (Fig. 1).

**Figure 1.**
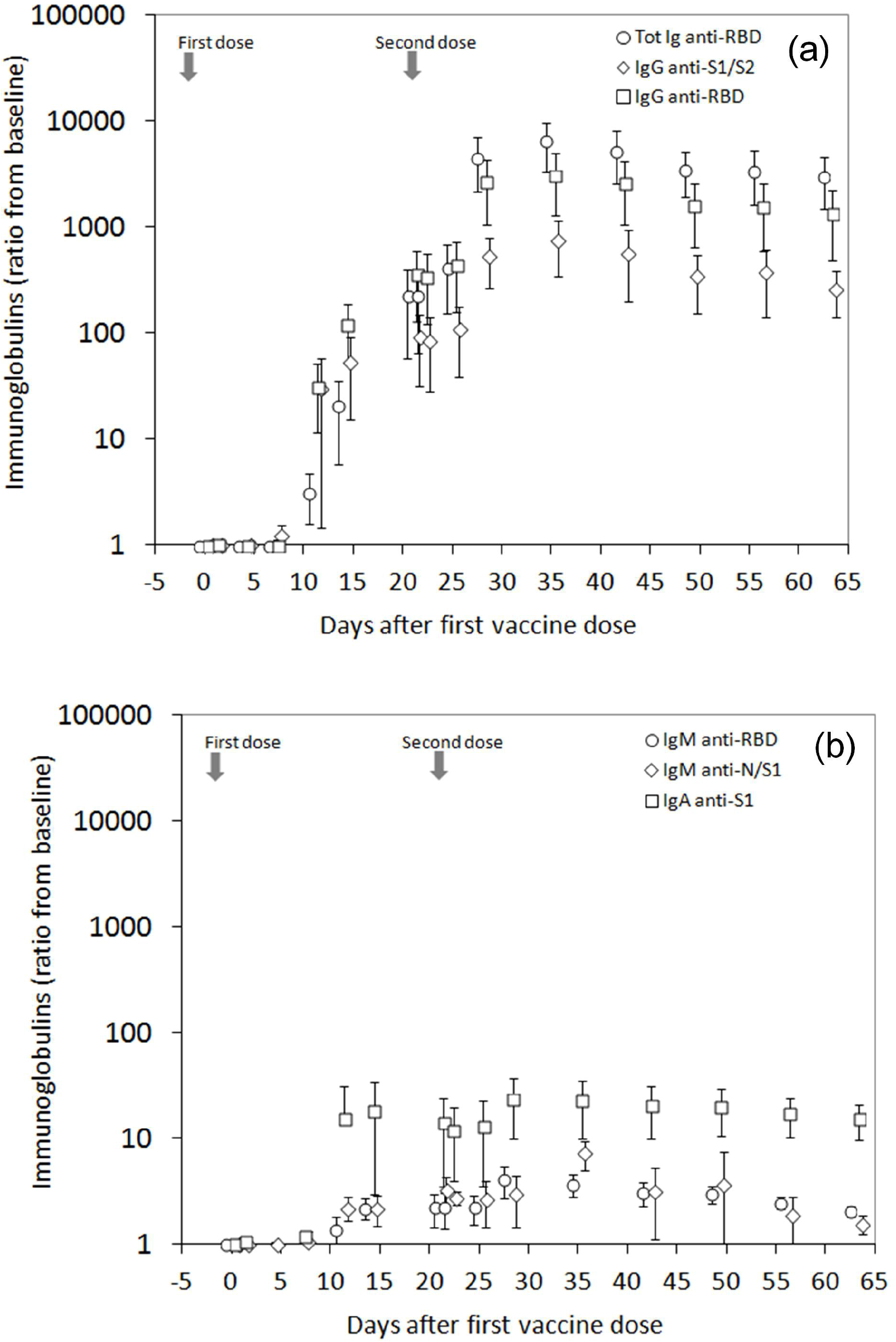
Overall kinetics of anti-SARS-CoV-2 (severe acute respiratory syndrome coronavirus 2) antibodies following BNT162b2 mRNA Covid-19 vaccination. Values are shown as mean ± standard deviation. *Ig, immunoglobulin; N, nucleocapsid; RBD, receptor binding domain; S, spike protein; Tot, total*

**Figure 2.**
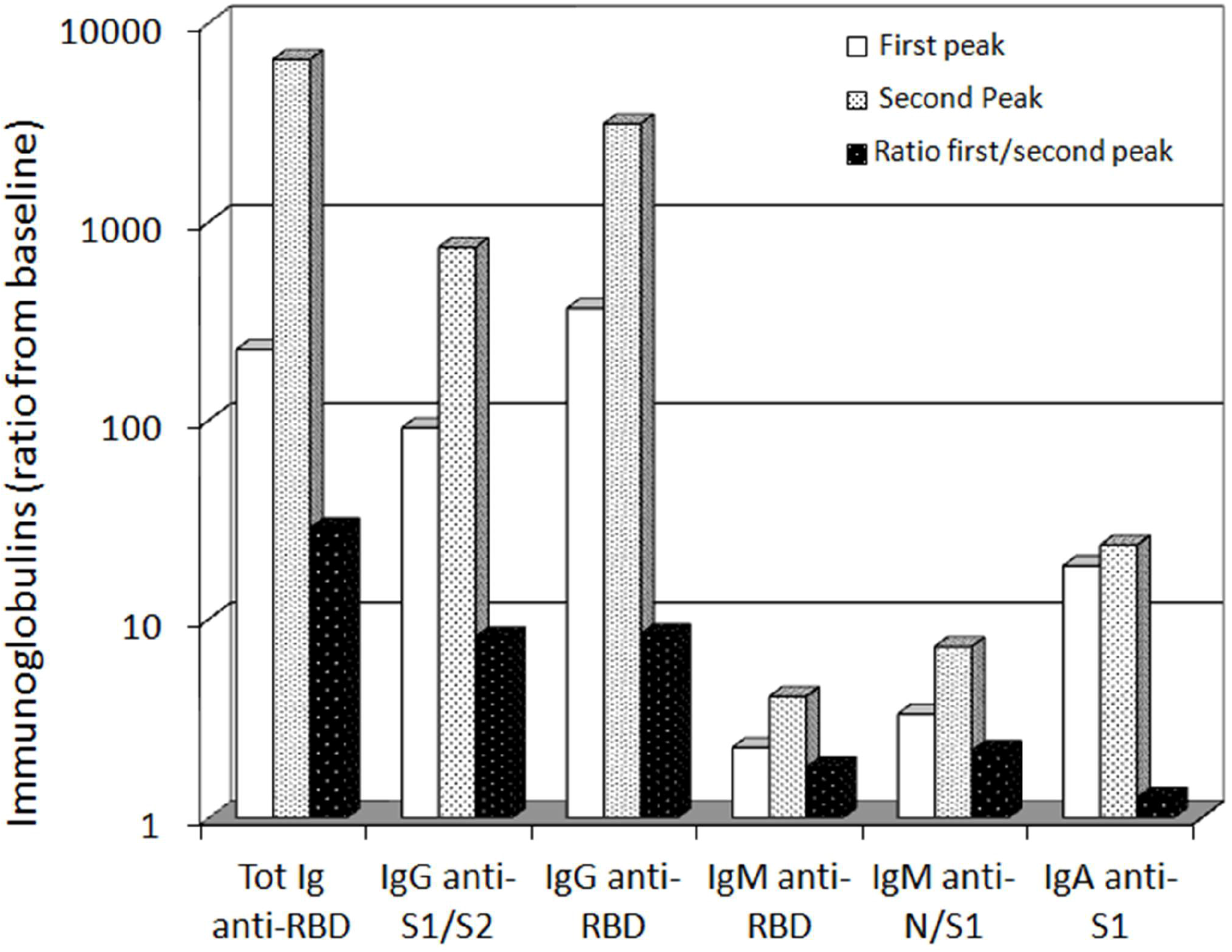
Peak values of anti-SARS-CoV-2 (severe acute respiratory syndrome coronavirus 2) antibodies observe after the first and second dose administration of BNT162b2 mRNA Covid-19 vaccine. *Ig, immunoglobulin; N, nucleocapsid; RBD, receptor binding domain; S, spike protein; Tot, total*

IgA anti-S1 level started to increase between 7-11 days after the first vaccine dose, but displayed a smooth decline between 14-21 days (Fig. 1b). The second vaccine boost was then effective to increase IgA anti-S1 levels by ∼1.3-fold compared to the first peak, reaching a second peak at day 28 (Fig. 2). The kinetics of IgA anti-S1 then displayed a slow and gradual decline, but the levels still remained over 15-fold higher than before vaccination.

The kinetics of IgM was very similar to that of the IgA anti-S1, though the IgM anti-RBD and IgM anti-N/S1 response with the immunoassays used in this study was substantially weaker (Fig. 1b), reaching the maximum peak after the second vaccine dose at day 28, which was only 4- and 7-fold higher than before vaccination (Fig. 2). Importantly, the level of both anti-RBD and anti-N/S1 IgM had declined to values similar to the baseline at 65 days after the first vaccine dose. The serum concentration of the spike protein, even doubling the sample volume in ELISA wells, remained undetectable (i.e., <1.35 ng/mL) throughout the study period with the manual immunoassay used in this study.

The Pearson’s correlations between the levels of different antibodies classes over time are shown in table 2. A highly significant inter-correlation was noted between total Ig anti-RBD, IgG anti-S1/S2 and IgG anti-RBD (all correlations r=0.99; p<0.001), whilst a lower but still good and significant correlation could be noted among the other anti-SARS-CoV-2 antibodies classes.

**Table 2.** Pearson’s correlation of anti-SARS-CoV-2 antibodies levels in three subjects vaccinated with BNT162b2 mRNA Covid-19.

| Antibodies | IgG anti-S1/S2 | IgG anti-RBD | IgM anti-RBD | IgM anti-N/S1 | IgA anti-S1 |
| --- | --- | --- | --- | --- | --- |
| **Tot Ig anti-RBD** | 0.99 (95% CI, 0.97-1.00)<br>p<0.001 | 0.99 (95% CI, 0.97-1.00)<br>p<0.001 | 0.88 (95% CI, 0.66-0.96)<br>p<0.001 | 0.69 (95% CI, 0.28-0.89)<br>p=0.004 | 0.75 (95% CI, 0.39-0.91)<br>p=0.001 |
| **IgG anti-S1/S2** | - | 0.99 (95% CI, 0.98-1.00)<br>p<0.001 | 0.92 (95% CI, 0.76-0.97)<br>p<0.001 | 0.76 (95% CI, 0.40-0.92)<br>p=0.001 | 0.79 (95% CI, 0.47-0.93)<br>p=0.001 |
| **IgG anti-RBD** | - | - | 0.92 (95% CI, 0.76-0.97)<br>p<0.001 | 0.71 (95% CI, 0.31-0.90)<br>p=0.003 | 0.78 (95% CI, 0.45-0.92)<br>p=0.001 |
| **IgM anti-RBD** | - | - | - | 0.78 (95% CI, 0.44-0.92)<br>p=0.001 | 0.93 (95% CI, 0.79-0.98)<br>p<0.001 |
| **IgM anti-N/S1** | - | - | - | - | 0.70 (95% CI, 0.30-0.89)<br>p=0.004 |

*95% CI, 95% confidence interval; Ig, immunoglobulin; N, nucleocapsid; RBD, receptor binding domain; S, spike protein; Tot, total*

## Discussion

Published information on immunogenicity of full-dose administration of BNT162b2 mRNA Covid-19 vaccine in real-world scenarios is still scarce and fragmentary. The humoral immune response after double 30 μg doses of Pfizer mRNA BNT162b2 Covid-19 vaccine was explored early on in macaques by Vogel et al. [10], who showed that a robust increase (i.e., ∼3 orders of magnitude) in anti-SARS-CoV-2 receptor biding domain (RBD) IgG could be observed 14 days after the first dose, whilst an even stronger response, exceeding 4 orders of magnitude from baseline, could be seen 7 days after the second dose. In a pilot trial, Krammer et al. longitudinally monitored the antibody response of 110 subjects receiving two mRNA COVID-19 vaccines (i.e., mRNA BNT162b2 and mRNA-1273) [11], reporting that most seronegative individuals developed an anti-SARS-CoV-2 spike protein IgG response (of ∼2 orders of magnitude compared to baseline) between 9-12 days after the first dose, increasing slightly over 3 orders of magnitude between 21-27 days. After the second vaccine dose, the anti-spike IgG antibody level further increased to approximately 3.5 orders of magnitude compared to baseline. Interestingly, the antibody response was anticipated and stronger in SARS-CoV-2 seropositive people. In another interesting studyrecently published by Wang et al., the authors showed that both of the two currently approved mRNA vaccines seem equally effective to elicit a sustained immune response, with production of antibodies of all classes, especially IgG, against SARS-CoV-2 spike protein and RBD at 2 months after the second vaccine dose [12]. Nonetheless, despite that the IgG anti-SARS-CoV-2 spike and RBD response was sizeable, exceeding 4 orders of magnitude from the baseline level, the other immunoglobulin classes behaved differently, with smaller increases and larger inter-individual variation. In particular, the levels of IgA and IgM were between 6-12 and 18-20 folds lower compared to that of IgG, whilst the intra-individual variation ranged between 26-30% for IgG, 103-105% for IgM and 59-65% for IgA, respectively.

Although the present report is limited to a 3-case series, it has many strengths compared to previously published studies assessing post-vaccination immune response. Specifically, we applied a narrow sequence of blood sampling, which allowed for (i) early identifcation and strict monitoring for the emergence and progression of anti-SARS-CoV-2 antibodies, (ii) the assessment of humoral response with measurement of all relevant antibody classes (i.e., total Ig, IgM, IgA and IgG), and (iii) the monitoring of the serum concentration of SARS-CoV-2 spike protein eventually produced after vaccination. This has hence allowed us to demonstrate that, in keeping with previous evidence, total Ig anti-RBD, IgG anti-S1/S2 and IgG anti-RBD displayed a sustained increase after the first vaccine dose, exhibiting an additional raise elicited by the second vaccine dose, after which a substantially stable plateau could be reached (Fig. 1a). This would provide a reasonable support to the recent findings of Chodcik et al, that a single mRNA BNT162b2 vaccine dose generates only ∼50% protection against severe COVID-19 illness [13].

Interestingly, an investigation following the Israeli nationwide vaccination plan, showed that a single vaccine dose had 46%, 57% and 62% efficiency for preventing SARS-CoV-2 infection, COVID-19 symptomatic illness and severe disease, respectively, but such efficacy increased up to 92%, 94% and 92% one week after the second boosting dose [14]. In the elderly population, which is more seriously affected by SARS-CoV-2 infection, Britton et al. found that martial mRNA vaccination (i.e., with a single dose) was only 63% effective to prevent COVID-19 in residents of skilled nursing facilities [15]. The coupled findings of lower neutralizing antibodies response and lesser real-life protection attainable after a single mRNA vaccine dose would hence suggest that stronger immunization should be pursued by a second administration, in order to boost immune protection, especially in the older population and against emerging SARS-CoV-2 variants [2].

Mirroring data garnered from the immune response seen after acute SARS-CoV-2 infections, the IgM response observed in this study was the weakest among all antibody classes, with the highest peaks of IgM anti-RBD and IgM anti-N/S1 observed at only between 4 and 7 folds higher than baseline (Fig. 2). This is in keeping with previous data published by Wang et al. [12], who also showed that the anti-RBD and anti-S IgM response after mRNA vaccination was 18-20 times lower than that of anti-RBD and anti-S IgM, thus corroborating general skepticism that this antibody class likely has limited importance for SARS-CoV-2 neutralization in vivo.

Importantly, we also provided the first evidence of a sustained (i.e., ∼15-fold) anti-SARS-CoV-2 IgA response elicited by BNT162b2 vaccine, still persisting for over 2 months after the first vaccine dose. Unlike other Ig classes, however, the response was flatter and the second vaccine dose only elicited a modest increase compared to the peak reached after the first vaccine dose (Fig. 2). Since the serum IgA value correlates strictly with that of their secretory counterpart [16], our findings would hence provide a reasonable background to the recent evidence that mRNA vaccines not only may be effective to consistently reduce the risk of developing severe COVID-19 illness, but could also confer important protection against SARS-CoV-2 infection [14]. In keeping with previous evidence, we also found that the relative increase of total Ig and IgG was 1-2 orders of magnitude greater than that of both IgM and IgA (Fig. 1) [12].

The spike protein serum concentration was undetectable (i.e., always lower than 1.35 ng/mL) with the method used in our study. This could be due to either a lack of ELISA antibody recognition of the recombinant form of SARS-CoV-2 spike protein produced after vaccination (despite consisting of full-length spike protein with two proline substitutions at K986P andV987P) [17], or due to cellular synthesis and release in the bloodstream of protein concentrations below the limit of detection of the assay. We cannot establish which of these two hypotheses may be true at this point in time, since BNT162b2 mRNA Covid-19 vaccine only contains mRNA encoding for SARS-CoV-2 spike protein and, therefore, does not react with ELISA antibodies targeting other epitopes present on mature proteins (e.g., the nucleocapsid). Nonetheless, these results are not really unexpected. Intramuscular injection of mRNA vaccines directly delivers nucleic acids into the muscle tissue, which contains an extensive network of blood vessels where many types of immune cells are present or can be rapidly recruited. Infiltrating antigen presenting cells (APCs), dendritic cells and even muscle cells themselves at the site of the injection are the main target of liposome-bearing RNA vaccines, along with other immune cells in the draining lymph nodes [18]. All these cells would hence uptake vaccine-bearing liposome particles, a process followed by cytoplasmic release of mRNA, which will then be translated into mature S (spike) protein, further expressed at the cell surface though major histocompatibility complex (MHC) I and II pathways [19]. Although it is theoretically conceivable that some amounts of endogenously generated SARS-CoV-2 spike protein could reach the bloodstream by direct exocytosis or intracellular release after disruption of muscle or immune cells membrane integrity, this concentration is perhaps too low to be quantified using conventional analytical techniques, such as ELISAs. Further studies, based on much more sensitive techniques, must be envisaged to elucidate this aspect.

It has now been clearly established that vaccination is a safe and effective option for averting the insurgence of a vast array of infectious diseases, and COVID-19 will make no exception [20]. The baseline assessment and long-term monitoring of humoral and (advisably also) cellular immune response before and after vaccination represent two mainstays of all healthcare policies aimed at preventing or containing the worldwide spread of SARS-CoV-2 infections and severe COVID-19 illness, as recently endorsed by national and international recommendations [21,22]. Although limited in size, our preliminary results have provided a reasonable background for defining the kinetics of different anti-SARS-CoV-2 antibody classes after mRNA COVID-19 vaccination, paving the way to larger and longer studies aimed to assess pan-humoral immune response and efficacy of different type of vaccines. Accurate anti-SARS-CoV-2 antibodies titration seems now unavoidable in many clinical circumstances [23], and will expectedly become even more important in an evolving scenario, characterized by occasional emergence of novel SARS-CoV-2 variants bearing specific mutations which may help evading immune recognition and response (i.e., B.1.351 and P1) [2]. Sustained neutralizing antibodies titers will hence be essential for maintaining a sufficient degree of protection against SARS-CoV-2 infection and illness caused by emerging new strains. To this end, monitoring anti-SARS-CoV-2 neutralizing antibodies (especially IgG and IgA) will help in defining whether additional strategies shall be envisaged, such as modifying the current vaccine formulations or revolutionizing administration plans, entailing for example the administration of additional vaccine boost(s) for enhancing neutralizing antibodies for a protective titer against new SARS-CoV-2 variants displaying potential for immune escape [24].

## Data Availability

Data will be available upon request to the corresponding author

## Research funding

None declared.

## Author contributions

All authors have accepted responsibility for the entire content of this manuscript and approved its submission.

## Informed consent

Informed consent was obtained from all individuals included in this study.

## Ethical approval

The study was reviewed and approved by the local Ethics Committee of Verona and Rovigo (2683CESC; February 16, 2021).

